# Causes of Autoimmune Psoriasis and Associated Cardiovascular Disease: Roles of Human Endogenous Retroviruses and Antihypertensive Drugs—A Systematic Review and Meta-analysis

**DOI:** 10.1101/2023.11.24.23298981

**Authors:** Aysa Rezabakhsh, Masoud H. Manjili, Hossein Hosseinifard, M. Reza Sadaie

**Affiliations:** Cardiovascular Research Center, Tabriz University of Medical Sciences, Tabriz, Iran; Department of Microbiology and Immunology, Massey Cancer Center, Virginia Commonwealth University, Richmond, VA, USA; Department of Biostatistics, School of Public Health, Hamadan University of Medical Sciences, Hamadan, Iran; NovoMed Consulting, Biomedical Sciences, Germantown, Maryland, USA

**Keywords:** Autoimmune disease, Psoriasis, Psoriatic cardiovascular disease, Human endogenous and nonendogenous retroviruses, Human immunodeficiency virus, Genome-wide genetic screening/virus variants, Drug-induced psoriasis, Biomarkers (biologic/immunologic/genetic)

## Abstract

Current treatments are ineffective to cure or prevent occurrences of autoimmune psoriasis and psoriatic cardiovascular disease/CVD. Psoriasis is associated with deregulated expressions of human endogenous retroviruses (ERVs) variants. ERV transcripts and proteins are detected in lesioned biopsies—without assembled viral particles—in addition to antibody and T-cell responses against ERV-K dUTPase. In persons living with HIV-1, manifestations of psoriasis are exacerbated variably. These may depend on multiple factors, differences in ERVs expressions, subtypes of HIV-1, and/or epigenetics. This article represents a quantitative risk assessment and meta-analysis approach with an attempt to assess causality. We surmise that mutated ERVs trigger aberrant proliferation and differentiation of keratinocytes, which in turn induce proinflammatory polarization. Independent risk factors and/or covariates with a range of relative risk/RR ratios appear to significantly impact the development of autoimmune psoriasis or immune intolerance, plausibly through ERVs genes activity. Given the antihypertensive drug’s potential in psoriasis development, a probable role in promising either ERVs activation or perturbations in epigenetic factors is questionable. Although the correlational nature of the data based on RR ratios prevents making robust conclusions, we reckon that the likelihood of attributable risk factors for certain antihypertensive drugs may stem from their pleiotropic effects or potentials for inducing ERV-mediated dysregulation of keratinocytes and/or endothelial cells. These findings expand our knowledge regarding ERV activations and HIV-1, antihypertensive drugs use, and incidents of psoriatic disease, and call for exploring cell-specific therapies aimed at blocking or reversing mutated ERVs gene activity toward attaining stable remissions in psoriasis and associated CVD.

## Introduction

Pathogenesis of human psoriatic skin lesions has never been associated with the presence of infectious virus-like particles. A multitude of dependent and independent variables, and risk factors, including mutated, nonreplicating, as well as replicating/particle-associated retroviruses, appear to exhibit relevant roles in the pathogenesis of autoimmune diseases and cancers ^1–3^. About 5-8% of the human genome contains endogenous retroviruses with sequences similarities to infectious human retroviruses, including type-C retroviruses, namely human endogenous retroviruses (ERVs) that are largely transcriptionally silent ^4^. Many ERVs consist of solitary transcription promoters namely long terminal repeats (LTRs) which can direct expression of nonviral genes – or partial/non-LTR sequences with long or short interspersed nuclear elements, some of which can express one or more viral structural (Gag, Pol/reverse transcriptase/RT, Env) and regulatory genes ^4^. ERVs are rarely transmissible, but are transmitted vertically in generations since dawn of the human evolution time ^1,4^.

Some ERVs exhibit various mutations, from single amino-acid exchanges to large truncated genes, deletions, and or insertions, exhibiting extensive heterogeneity, and polymorphisms. Many of these alterations are localized in conserved exons or coding sequences while others are distributed in noncoding introns or LTR promoters which generate RNAs from enclosing genes ^5–8^. In psoriasis, certain variants and mutants of ERVs may be associated with the initiation, progression, and severity of the disease, and possibly psoriatic cardiovascular disease/CVD (see further).

Previous studies have largely demonstrated associations between activations of immune cells, the release of inflammation-inducing cytokines, and interleukins from epidermal dendritic/Langerhans cells, and shared genetic risk factors in relation to ERVs, as potential causes for uncontrolled proliferation of dermal keratinocytes in psoriasis and psoriatic CVD ^8–10^. It remains unclear whether the immune intolerance and induction of inflammation are routed in transcriptional activities of ERV genes partly to suppression of protective genes in keratinocytes and partly to aberrant activation of immune cells are consequences for triggering autoimmune psoriasis. Furthermore, real reasons for incidents of other disease burdens in psoriasis, in particular psoriatic CVD remain unknown.

### Hypothesis

T cells are present in normal skin to provide immune surveillance without causing any harm. They are also involved in psoriasis, perhaps owing to conditions resulting in intolerance of the immune response in the skin. The cause(s) of tissue-specific immune intolerance in psoriasis and PsD are currently unknown. It is conceivable that ERVs trigger and establish uncontrolled proliferation and differentiation of keratinocytes and the release of inflammation-mediating chemokines that in turn induce cytokines/interleukins in migrating epidermal immune cells— simultaneously or in a sequential-temporal manner—in the effect site and concurrent developments of psoriasis and CVD. We hypothesize that tissue-specific mechanisms involving the mutated ERVs (expressions), rather than the chronic systemic inflammation, precede inflammatory immune responses in the skin and are the contributory causes of psoriasis and psoriatic CVD.

Plausible medication-induced psoriasis might also be rooted in a drug compound-induced activation of ERV elements—involving LTR promotor or other elements such as demethylation or inhibition of histone deacetylase in chromatin—may also result in activations of the ERVs. Taken together, these events can induce perpetual immune intolerance and inflammation in psoriasis and psoriatic CVD, as consequences of releasing sequestered cellular autoantigens from keratinocytes. In that sense, multiple ERV variants (e.g., -E, -K, -W, -9) might trigger and maintain psoriasis, and concurrently “trans-disease” symptoms, such as psoriatic CVD. Furthermore, vascular endothelial tissue presumably harbors chemical-inducible “high-risk” ERV genes, along with keratinocytes and epidermal immune cells (dendritic/Langerhans-like cells). Normal cellular genes are overactivated or disrupted, presumably through transcriptional and/or post-transcriptional dysregulation of ERV genes.

### Evaluating the hypothesis

In this article, plausible causal agents and shared links between distinct ERVs and related transcriptional activations and suppression of ERV-related genes as well as ERV-directed gene expression, from the perspective of dependent variables, are researched and discussed. Published datasets from cells in culture to genome-wide screening to clinical outcome measures using liquid/tissue biopsies biomarker assays are summarized, tabulated, and evaluated, by a risk assessment approach and trans-disease meta-analysis, in an attempt to determine whether datasets support the idea of ERVs as potential root causes of psoriasis and psoriatic CVD. In addition, datasets in published literature are reviewed and relevant information are quantitatively analyzed and discussed.

### Search Strategy

Scopus, PubMed Central, Medline, the NIH National Library of Medicine, Google Scholar databases, and preprint servers (medRxiv) were searched using the listed keywords up to June 2023 resulting from Medical Subject Headings (MeSH): “human endogenous retrovirus” OR “human ERV” OR “hERV” OR “human endogenous retroviruses” OR “human ERVs” OR “hERV” OR “hERVs” OR “HERV” OR “HERVs” OR “HERV-k” OR “HERV-H”

AND

“Psoriasis” OR “PSORS1C2 protein, human” OR “CARD14 protein, human” OR “FABP5 protein, human” OR “PRINS RNA, human” OR “PSORS1C3 lncRNA, human” OR “PSORS1C1 protein”

AND

“Antihypertensive Agents” OR “ANTIHYPERTENSIVE AGENTS” OR “DIURETICS” OR “human endogenous retroviruses” OR “calcium-blocking agent” OR “ANGIOTENSIN II TYPE I RECEPTOR ANTAGONIST” OR “beta-1-adrenergic antagonist” OR “angiotensin-converting enzyme inhibitor” OR “BETA-ANTAGONISTS” OR “ADRENERGIC ALPHA-ANTAGONISTS” OR “ANGIOTENSIN-CONVERTING ENZYME INHIBITORS” OR “GANGLIONIC BLOCKERS” OR “CALCIUM CHANNEL BLOCKERS”

AND

“systematic review” OR “review systematic” OR “systematic review” OR “ review” OR “meta-analysis” OR “analysis, meta” OR “meta analysis”

The publication date was used as unfiltered and recent ‘cited by’ or associated publications were also surveyed. The articles that were relevant to the scope of this article—including all the main evidence, original publications, cohorts and clinical studies, case reports and some review articles, alongside the systematic reviews and meta-analysis—were considered, reviewed and cited. Records of the search and review approach for the key articles are depicted in **Figure 1**. Other clinical manifestations of psoriasis, such as psoriatic arthritis/PsA and infection-associated psoriasis other than HIV-1, were largely excluded from the review. In addition, we took an integrative umbrella review approach, which includes review and analysis of the basic science empirical data, disease models and clinical reports, centered on the study hypothesis. It entails narratives and quantitative assessments, tabulations, summations of the datasets and illustrations of the relevant descriptions. This approach in part adopted a previous technique recommending to contextualize information when searching for putative risk factors, etiology, and or sources of risk, such as hazard exposure susceptibility, to attain unbiased data extractions based on the totality of evidence ^11^.

**Figure 1.**
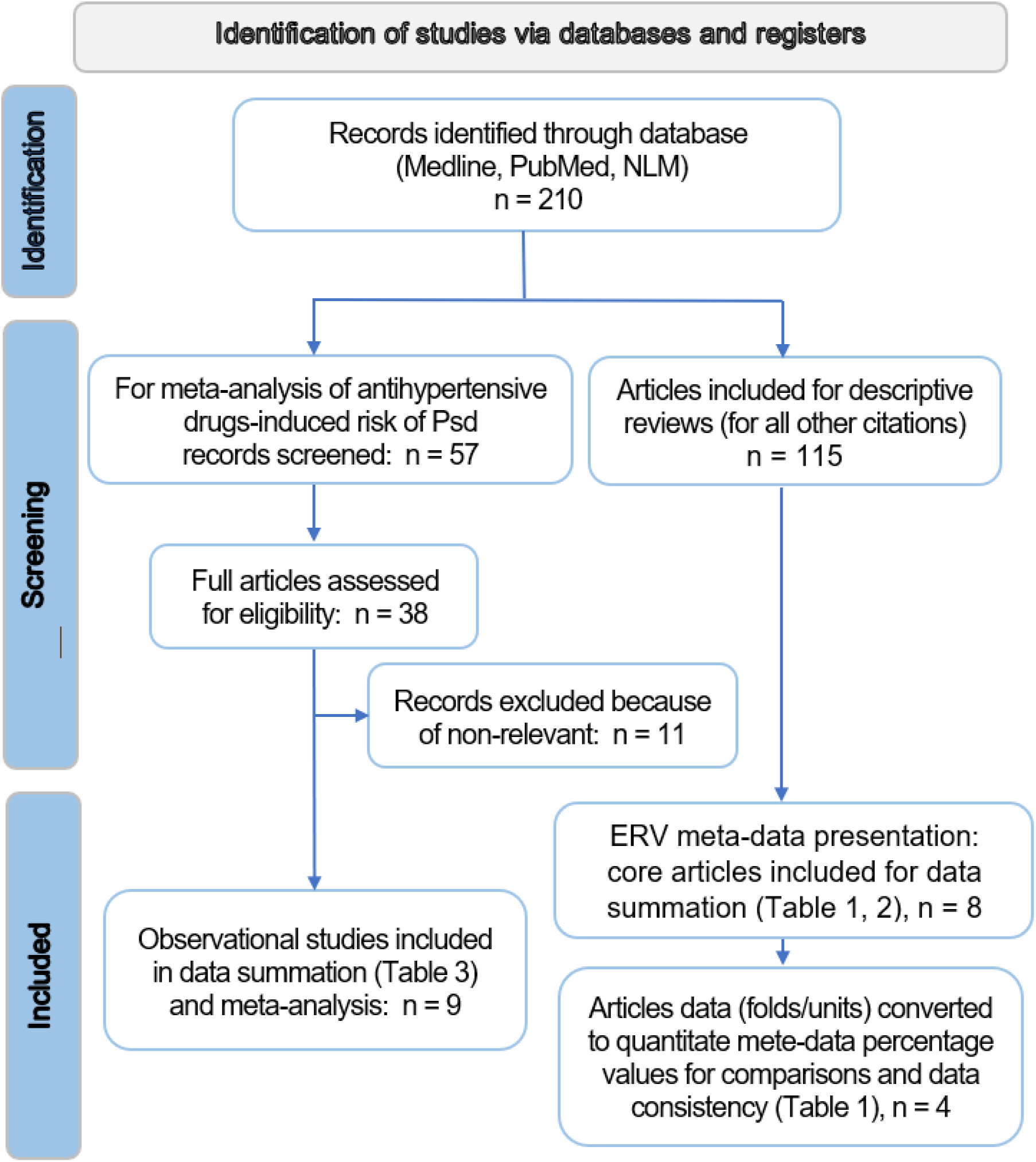
The search strategy flowchart

### Meta-analysis

Multiple studies have previously reported the likelihood of associations between antihypertensive drugs and incidents of psoriatic CVD. Literature search revealed ten eligible studies reporting odds ratios, without determining relative risk and the magnitude of risk difference between different studies. We adopted a meta-analysis approach to overcome limitations of the interpretations in these studies by calculating the relative risk/RR, risk difference between study groups and class of medications—as a foundation toward determining the number needed to treat/NNT values for additional insights in clinical investigations in the future. The points of estimates, odds ratios/OR values with 95% confidence interval (CI) were extracted from the articles, and then, given the somewhat lower prevalence of antihypertensive drugs-induced PsD (≤ 10%), it enabled to convert the odds ratios into the relative risk/RR ratios, as described ^12^ for better interpretability of the risk of PsD amongst this subset of patients. Inhomogeneity between studies was calculated using I-square (I^2^) and Q-value statistics, in which, the I^2^ value >50% was considered the presence of heterogeneity in the studies. Using a random effect model, the RR values of the included articles were merged and finally the pooled RR value was estimated. Analysis was performed in the subgroups, in accordance with drug classes, study design, risk stratification and disease type. A probability value of less than 5% was considered a significant level. The Comprehensive Meta-Analysis (CMA) software was used for these calculations. The multiple datasets were then tabulated and compared.

## Published Research

Psoriasis has been proposed to be associated with the expression of ERVs for the induction of inflammatory immune responses in the skin. The data are inconclusive because of some reports fail to show transcriptional activation as the only correlation in skin lesions. In addition, analysis of PBMCs from 69 patients with psoriasis revealed decreased expression of ERV-E compared with its expression in 20 healthy controls ^13^. On the other hand, UV light induced the expression of ERV in keratinocytes, and in turn, promoted the production of proinflammatory cytokines by keratinocytes and their proliferation in an autocrine fashion involving IL-6 and IL-22 ^14,15^. Such paradoxical observations can be explained when keratinocyte-derived cytokines are differentiated from those produced by inflammatory immune cells. While the former may be triggered by ERV expression in keratinocytes, the latter could target immunogenic components of ERVs and result in the inhibition of ERVs.

Genome-wide association, RT-PCR gene expression, and bioinformatics studies have identified numerous polymorphisms and rampant integrations of ERVs. In fact, different ERVs are overexpressed in cancer and autoimmune disease, and can also either disrupt or enhance normal expression of human genes, via integration of the viral promoter coding sequences, hence mediating transcriptional activation or suppression of the cellular genes. Remnants of the functional intragenic ERVs largely consist of LTR segments and lack important viral structural open reading frames (ORFs), capsid/envelope/polymerase (RT, protease, integrase), and regulatory genes. Functional single LTRs can act as promoters/enhancers to direct expressions of a variety of human genes ^5^. For instance, up to 50 copies of the ERV-E type are integrated within multiple different genetic loci in human chromosomes, which mostly exhibit point mutations, deletions or truncations ^16^.

ERVs, -E, -K, and -W are particularly interesting as a subset of these variants exhibits expressions of at least one viral variant structural genes, including *pol*, *gag*, and *env*, resulting in a readily detectable reverse transcriptase/RT, Env and Gag proteins in healthy tissue while significantly either accumulate are suppressed in distinct pathological conditions ^2,8,17–20^. Unlike the expression of ERV-E in skin cells, PBMCs of psoriatic patients contain much reduced expressions of ERV-E transcripts compared to healthy persons ^13^ (see further).

## Psoriatic Cardiovascular Disease/CVD

### Risk Factors Involved in Psoriasis-induced CVD

Psoriasis disease (PsD) is an autoimmune, chronic, and systemic inflammatory disease, affecting 2-3% of the U.S. population ^21^. The severe form of PsD is more likely to confer between 25-50% of increased relative risk of CVD, which has a positive correlation with skin severity ^22^. Available evidence established an increasing prevalence of CVD in patients with PsD with higher morbidity and mortality worldwide.

Guidelines issued by the American Academy of Dermatology/National Psoriasis Foundation (NPF) and the American Heart Association/American College of Cardiology (AHA/ACC) acknowledged that beyond the skin and joints involvement, psoriatic-induced CVD, e.g., myocardial infarction and stroke, as well as related premature mortality are concerning health issues in which prevention measures should be taken into account ^23,24^. Furthermore, it has been assumed that the current state of knowledge is in favor of genetic links between PsD and CV risk factors, consisting of shared genetic background, gene–environment crosslink, or epigenetic alterations ^25^.

Noticeably, it has been estimated that the incidence rate of both CVD and major adverse cardiovascular events (MACE) was slightly higher in psoriasis arthritis (PsA) when compared with PsD ^26^. However, the results of a meta-analysis revealed that following systemic therapy in patients with PsD and/or PsA, the risk of all cardiovascular morbidity and mortality significantly decreased [relative risk (RR), 0.75; 95% confidence interval (CI) 0.63 to 0.91; *p* = 0.003)] ^27^.

Albeit, established nontraditional and traditional risk factors have a substantial role in CVD progression, PsD exclusively provides an excessive inflammatory response as an independent and additional cardiovascular risk factor ^28,29^. In this respect, underlying inflammation response can play a key role in atherosclerosis progression ^30^, implying an overlapping mechanism with PsD. Also, the international classification of disease-9 codes, defined as billable medical codes, is practical for identifying CVD and the associated risk factors ^31^.

In a cohort of 66,420 women with hospital-diagnosed PsD, the odd ratios of traditional/nontraditional cardiovascular risk factors in whom were at higher risk of heart failure and ischemic cerebrovascular disease were assessed ^32^. The results of this study indicate a gender-specific possibility of CVD prevalence among PsD patients, which needs further investigation to prove this finding. Notably, a chronic stress-related neural activity, named amygdalar activity, could substantially contribute to increased subclinical CVD risk in PsD patients ^33^. Noteworthy, the gut composition differs between individuals with and without CVD, suggesting that gut microbiome dysbiosis might be associated with an incremental risk of CV events in psoriatic patients, which can be retrieved by the positive impacts of biologic therapies ^34^. Based on the pooled analysis of a recent meta-analysis, it has been also emphasized that PsD can be categorized as an independent risk factor, which is associated with the progression of adverse CV outcomes, including myocardial infarction, stroke, arrhythmia, ischemic heart disease, thromboembolism, and CV death, particularly in moderate-to-severe patients ^35^.

### The Shared Immunopathogenic Pathway

It has been inferred that there is an immune-based cross-talk between PsD and cardiovascular comorbidity. At the cellular level, over-activation of T cell subsets, myeloid cells, activated platelet, up-regulation of interferons (IFNs), and pro-inflammatory mediators accompanied by various cytokines, including interleukins (IL)-6, IL-17, IL-12/23, nitric oxide, and TNF-α are predominately implicated in the pathogenesis of psoriasis ^21,36,37^.

Following inflammatory disorders, three main events can occur in cardiovascular status, including fast-tracked coronary atherosclerosis, atrial fibrillation, and ventricular myopathy with a preserved ejection fraction ^38^. In view of the pathophysiology attributed to this comorbidity, the presence of inflammatory pathways, secretion and release of adipokines, oxidative stress, microparticles (miRNAs), insulin resistance, angiogenesis, and coagulopathy have been proven to play crucial roles ^39^.

In both psoriasis and vascular atherosclerotic lesions, the levels of macrophage, T helper 1 (Th1), and Th17 are increased. Simultaneously, Th1 lymphocytes induce antigen-presenting cells, Th1-related cytokines, and other T cells. The overexpression of the cytokines, in turn, could develop atherosclerotic plaques, as well as CVD. On the other hand, existing loss-of-function regulatory T cells contribute to diminishing the anti-inflammatory responses provoked during psoriasis and atherosclerosis ^40^. Consequently, the possible cross-talk between the presence of atherosclerotic and psoriatic plaques is partially referred to a shared inflammatory pathway mainly mediated by Th cells, aberrant angiogenesis, and endothelial dysfunction, as well as reduced level of dynamic endothelial progenitor cells ^41^.

It is worth noting that the dynamic levels of angiotensin-converting enzyme (ACE), renin, and endothelin-1 (ET-1) are elevated in PsD patients resulting in higher blood pressure. This axis by promoting T-cell proliferation consequently develops the inflammatory process of atherosclerosis ^42^. Besides, long-term exposure to systemic inflammation in severe PsD can lead to coronary microvascular dysfunction and vasculitis, at least in part, because of the increased levels of circulating neutrophils ^22^.

In addition, the role of microRNAs, as well-known small, single-stranded, noncoding RNA molecules with a crucial role in gene expression, in psoriasis-induced CVD has also been explored ^43^. For example, the expression of miR-200c is increased in PsD patients, reflecting endothelial dysfunction. In addition, a composite of miR-200c + miR-33a+ miR-33b by a negative impact on cholesterol homeostasis, resulting in atherosclerotic plaque destabilization ^44^. Among all identified miRNAs, miR-22 has represented a dual effect. In better words, the miR-22 expression is elevated in cardiomyocytes hypertrophy, while down-regulation of miR-22 is observed in the presence of atherosclerotic plaques ^44^. Also, the overexpression of miR-378a and miR-340 has been reported to develop CV events in PsD patients ^45^. Although the utility of these microRNAs as standard diagnostics in human laboratory medicine has yet to be proven, their involvements with the deregulated transcriptions of the ERVs in the posttranslational ‘gain-of-functions’ of ERVs RNAs and proteins (e.g., dUTPase, RTs, Gag, Env) are unexplored.

### The Panel of Biomarkers

Given that PsD is termed an inflammatory overload disease, remarkably higher serum levels of C-reactive protein (CRP) and IL-6 would be reliable predictors of CVD risk among PsD patients ^46^. Besides, dyslipidemia, especially increased oxidized low-density lipoprotein (ox-LDL) levels, with pro-inflammatory and pro-atherogenic potential, followed by two prominent adipokines (i.e., adiponectin and leptin) are considered other laboratory values to assess CV status in these patients. A recent cohort study also found an association between dynamic changes in cardiac biomarkers such as cardiac troponin I (cTnI) and N-terminal pro-brain natriuretic peptide (NT-proBNP) which may indicate the excess risk of clinical CV events in PsD patients independent of traditional risk factors (such as hyperlipidemia, hypertension, smoking, and obesity) ^47^. Besides, red cell distribution width (RDW) and mean platelet volume (MPV), as feasible and cost-effective laboratory tests, were also introduced as prognostic indicators of MACE, through the identification of bone marrow and plasma thrombogenicity which can be employed as advantageous aids in clinical making-decisions ^48^. Interestingly, the prognostic potential of urinary orosomucoid (u-ORM), a novel inflammatory biomarker, alongside the u-ORM/u-creatinine ratio for low-grade inflammation can be applied as an indicator of CV risk in patients with mild-to-moderate PsD ^49^.

### Biological Treatment

From a scientific point of view, biologics targeting TNF-α, IL-12/23, and IL-17 (particularly IL-17A) are recommended potential therapies that can improve both PsD and atherosclerosis by modulating the inflammatory response, highlighting an integrated therapeutic strategy ^22,50,51^ however to reach a stable remission appears unrealistic as such therapies target the symptoms rather than the cause(s). In a cohort study, TNF antagonists exerted a favorable efficacy against CV risk in PsD patients with appreciable safety in PsD patients considering probability of 50%, response time, 90% improvement, probability of American College of Rheumatology (ACR) 20 response, sustainability of success, and probability of mild and severe adverse events (AE) ^52^. Ustekinumab, a monoclonal antibody used for Crohn’s disease, ulcerative colitis, plaque psoriasis, and psoriatic arthritis, has the potential to modulate serum levels of inflammatory and CV risk proteins, including E-selectin, Myeloperoxidase, von Willebrand factor, C-X-C motif chemokine 10, Cystatin-B, TNF receptor 1, NT-proBNP, IL-8, and Urokinase plasminogen activator surface receptor, but IL-6, in PsD ^53^. However, it should be noted that some medications prescribed for patients with PsD may exert worsen effects in terms of CV risk.

Considering this scenario, a recent post-hoc analysis of various phases of clinical trial study reported that tofacitinib exposure, a Janus kinase inhibitor used for PsA treatment, increase the incidence rate (IR, with 95% CI) of major adverse cardiovascular event/MACE in whom had a baseline history of atherosclerotic CVD (ASCVD) [0.75 (0.02-4.21)], followed by the high (≥ 20%) 10-year risk of ASCVD [1.37 (0.03-7.63)], and the high risk of malignancies [1.13 (0.45-2.33)] in a subset of PsA patients [56]. The highest IR of MACE with a history of ASCVD [1.87 (0.69-4.08)], a high 10-year risk of ASCVD [1.49 (0.55-3.24)], followed by the high risk of malignancy [3.48 (1.90-5.83)] was reported in the PsD patients ^54^.

In addition, several biological disease-modifying antirheumatic drugs (bDMARD), including IL-12/23 and IL-17 inhibitors prescribed in PsD patients due to anti-inflammatory impacts led to enhance the risk of CV adverse events, such as acute myocardial infarction and ischemic stroke (IL-12/23: hazard ratio/HR 2.0, 95% CI 1.3-3.0; IL-17: HR 1.9, 95% CI 1.2-3.0) particularly in new users, compared with TNF-α antagonists ^55^. While, for arterial intima-media thickness (IMT) observed in severe PsD patients, IL-17 inhibitor therapy is introduced as a promising therapeutic option, due to long-term benefits through exerting an anti-inflammatory effect on vascular beds following vascular events i.e., vasculitis ^56^.

## Possible Causes

In general, ERVs can provide a variety of pathogenic elements, like RNA: DNA duplexes, coding and noncoding transcripts, transposable intergenic insertions or recombination, as well as proteins and even some virus-like particles, all of which can interfere with gene expression in normal somatic, immune and cancer cells, and conversely provide a protective role ^5,8,57^. These ERV elements are presumably involved in inducing aberrant proliferation and differentiations of keratinocytes and can act either as autoantigens (through molecular structure(s) mimicry mechanisms) or superantigens or both generate systemic polyclonal T-cell activations and inflammation. Roles of regulatory noncoding small molecule RNAs that are interrelated to ERV LTRs promoter activity and intergenic sequences are largely unexplored ^58^.

Recently, genome-wide RNA sequencing and differential screenings demonstrate that numerous ERV-K genes are transcriptionally suppressed, whereas only a small subset is upregulated in lesional and nonlesional psoriasis ^7^. The authors suggest that proinflammatory environment results in a general suppression of ERV transcripts, and further point out that crucial other intertwined mechanisms, including chromatin modifications, locus-specific effects of cis-/trans-regulatory elements, in addition to immunoreactivity of functional ERV proteins should be explored ^7^. Conversely, initiation of psoriasis might also be rooted in the suppression of cell-cycle inhibitory checkpoints triggered by abortive transcription of LTR-directed coding sequences that otherwise play a part in the normal physiology of keratinocytes.

### Human Endogenous Retroviruses Activation and HIV-1

Among human endogenous retroviruses, several variants (ERV-E/-K/-W/-9) are transcriptionally and translationally highly activated or altered in expressions, as quantitated using qRT-PCR and protein assays, although assembly of virus-like ERV particles is undetected in psoriasis. Detection of transcripts signals of ERV-K/-9 are demonstrated using primers and peptides representing ERV-K (env, gag, pol, rec) and ERV-9 genes in skin biopsies in qRT-PCR and serology assays ^20^. While the expressions of ERV-E,-W/-09 are increased in skin samples of psoriatic patients ^9,19^, transcripts of all major ERV-K proteins are substantially decreased compared to healthy persons ^20^ (**Table 1**).

**Table 1.**
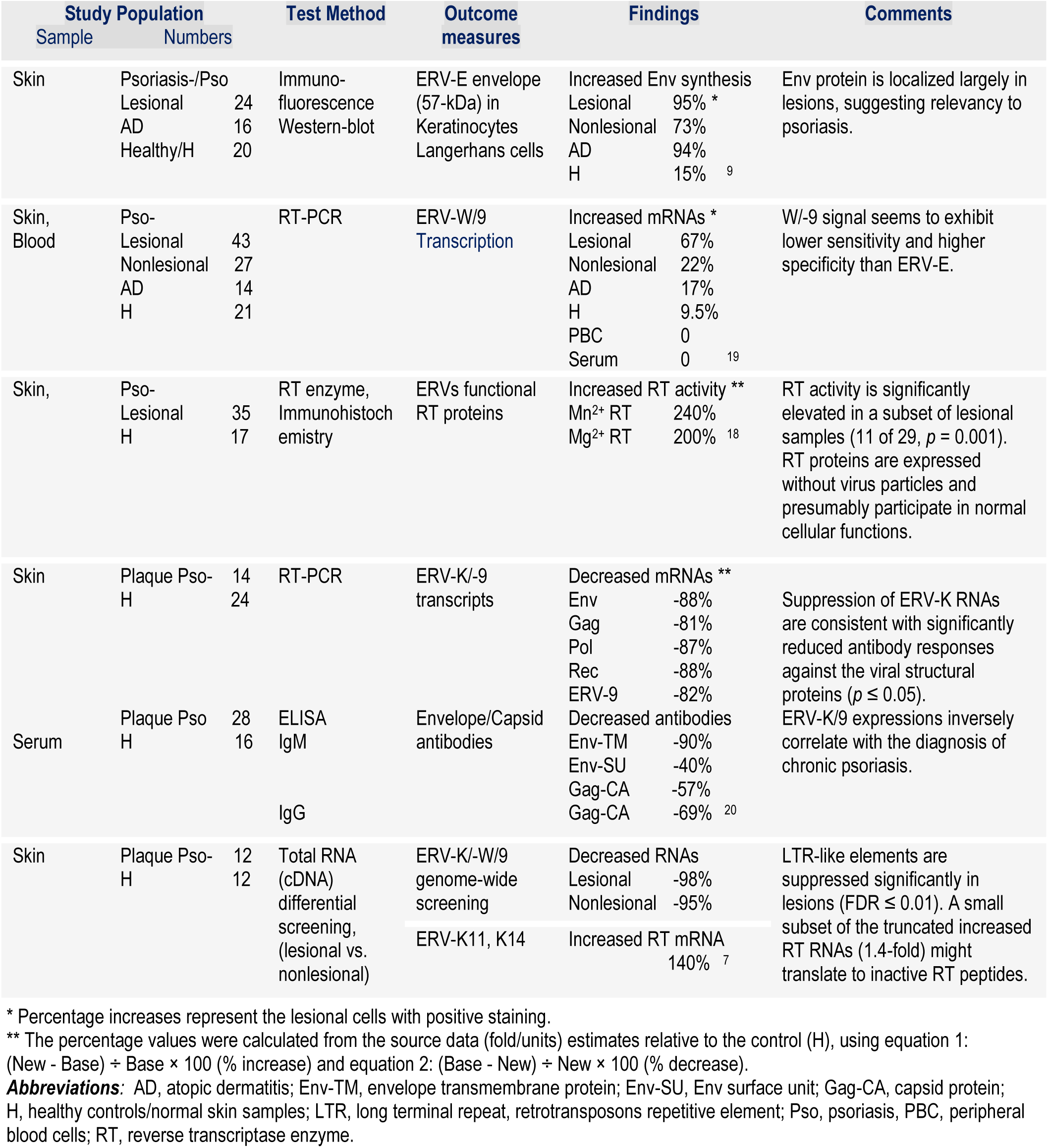
Gene expression evaluations of ERV variants using liquid/tissue biopsies.

Laboratory examination of biopsies from a cohort of patients with psoriasis, atopic dermatitis, and healthy persons as control arm, demonstrates that ERV-E Env protein is expressed in dermal T-cells infiltrates CD3+/CD4+ in both perilesional (16 of 22, 73%) and nonlesional psoriatic skin, and in CD8+ and CD1a+ Langerhans cells, using immunofluorescent staining ^9^. The same protein appears present in lesional atopic dermatitis/AD (15 of 16, 94%) localized in cell nucleus. Although nonspecific staining was ruled out, the role of this autoantigenic protein is unclear. Moreover, considering that the reaction was largely detected in perilesional and lesioned skin tissue, but nonregional biopsies, the authors conclude that ERV-E activity may be relevant to the disease pathogenesis. In this observation, multiple additional peptides were detected in Western blots maybe because polyclonal antibodies were utilized. The positive reactions could represent metabolic byproducts of the Env protein or cross-reactivity with unrelated autoantigens with shared epitopes.

Consistent with this, an increased synthesis or accumulation of envelope protein of ERV-E was found in psoriatic lesions, including keratinocytes, CD3+/CD4+ lymphocytes, but CD8+ and Langerhans-like cells ^9^. Transcripts of these, however, were not detected in peripheral blood mononuclear cells (PBMCs), presumably downregulated as a result of viral defense mechanisms ^13^. Functional RT protein (in the absence of viral particles) is also significantly higher in lesional samples than those from healthy persons, in protein extracts from biopsies, observed only in a subset of samples using immunochemistry ^18^. Whether RT and Env proteins trigger and or accelerate the proliferation and conversely differentiation of keratinocytes that result in apoptosis, the release of autoantigens, and inflammation is unexplored.

Notably, while expressions of ERV-E,-W/-09 are increased in skin samples of psoriatic patients ^9,19^, genomic transcripts of all major ERV-K proteins are significantly suppressed in samples from psoriasis compared to healthy controls ^20^. Recently, genome-wide differential screening of total RNA extracted from biopsies demonstrates that only a small subset of ERV-K RNAs, including truncated RT enzyme, are up-regulated in lesional tissue, majority of RNAs representing LTR retroelements are down-regulated in psoriasis (**Table 1**) ^7^.

While there is no indication that ERVs cause psoriasis, compelling findings from a large case-control study, with a mean age onset of 24.6 years, suggest that five common ERV-K dUTPase variants are significantly associated with psoriasis, with the strongest at the missense mutation K158R. Elevated autoantibodies were also detected against recombinant wild-type ERV-K dUTPase in addition to higher T-cell responses ^8^. The authors assert that human dUTPase might have other roles like DNA-binding protein or interact with peroxisome proliferator-activated receptor (PPAR) isoforms like PPAR delta, previously implicated in psoriasis. Another mutant, for instance 155G/158R, previously termed the ‘high-risk’ variant is also the most common haplotype in psoriasis ^17^ (**Table 2**).

**Table 2.** ERV mutants in psoriatic lesions: case-control diagnostics and experimental laboratory studies.

| Target gene | Study Population<br>(No. of Samples) | Outcome Measures | OR<br>[95% CI] | Findings | Comments |
| --- | --- | --- | --- | --- | --- |
| ERV-K dUTPase e.g., K158R 'high-risk' variant | Biopsies<br>Pso- 708<br>H 349 | DNA sequence, Antibody | 2.36 [1.91–2.92]<br>$p = 3.28 \times 10^{-15}$ | Increased expression of five common ERV-K mutants correlates with elevated antibody responses against ERV dUTPase. K15R mutant is highly positively associated with psoriasis<sup>8</sup>. | ERV-K mutants exhibit variable degrees of proinflammatory signals, from 'low- to high-risk'. Expression of some mutants positively whereas others negatively associated with psoriasis. Further research is needed to determine the consistency of associations in predefined inclusion criteria. |
| Multiple mutants, e.g., 155G/158R, Δ142-171 | Primary hDCs, Keratinocytes in Culture | Functional studies on proteins, Immune biomarkers | - | Increased TNF-α, IFN-γ, and TLR-2 dependent activation of NF-κB (4.5 to 31.5-fold), and chemokines (CCL-20/23, RANTES), interleukins (IL-12/p40, IL-1β/IL-8/-10/17/-23), more in hDC than keratinocytes and trigger Th1/Th17 response, while some mutants inhibit, independent of enzymatic activity<sup>59</sup>. | |
| 155G/158R | Skin, Blood<br>Pso- 4<br>H 4 | Haplotype sharing analysis | - | Confirmed mutants are detectable in Pso skin and blood and located in the psoriasis susceptibility gene locus<sup>17</sup>. |  |

**Abbreviations:** dUTPase, deoxyuridine triphosphate nucleotidohydrolase; hDCs, human primary dendritic cells; OR, odds ratio; Pso, psoriasis; RT, reverse transcriptase enzyme; TLR-2, toll-like receptor 2; K158R/GR, lysine (K) to arginine (R) or glutamine (G) to arginine conversions.

The presence of autoantibodies against the mutant epitopes, as well as other mutated motifs, remain also unexplored. However, in support of the high signal detection and functional studies on the relevance of dUTPase mutants in initiation of immune intolerance, emphasis is put partly on positive T cell reactivity, and the observation that dUTPase variants stimulate cytokine responses in primary dendritic cells in culture ^59^. Taken together, potential causal relationships between mutated ERVs expression and pathogenesis of keratinocytes and induction of proinflammatory polarization are illustrated in **Figure 2**.

**Figure 2.**
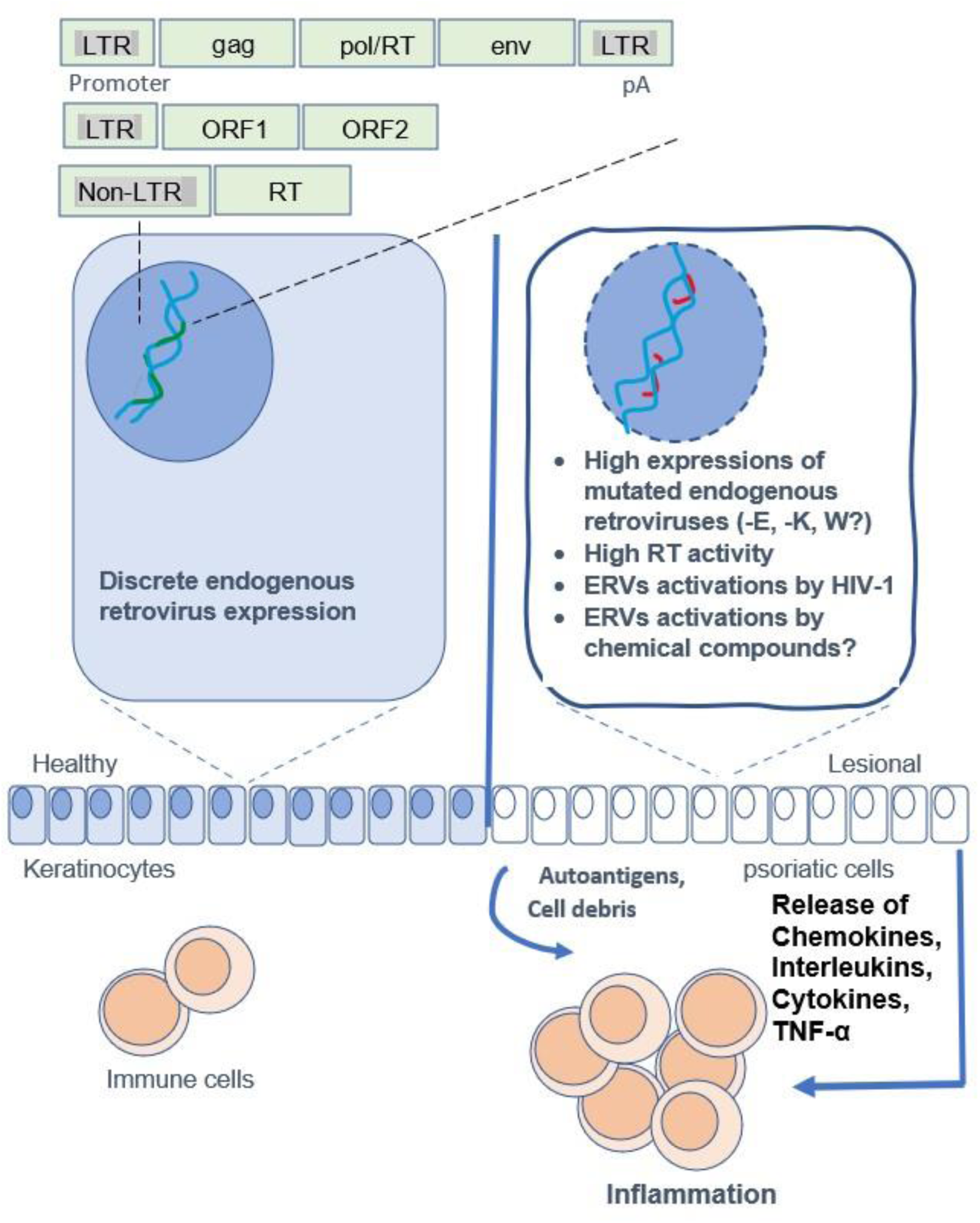
Schematic representation of human ERVs gene expressions in associations with induction of psoriatic lesion. Elevated expressions of the mutated ERVs presumably trigger aberrant proliferation and differentiation and promote cytopathic effects, resulting in programmed cell death/apoptosis in keratinocytes, which can release cell debris and autoantigens in epidermal tissues, and result in mobilization and expansion of resident immune cells and inflammation. ***Abbreviations***: LTR, long terminal repeat/ERV promoter, ORF, open reading frames, gag/pol/RT/env, capsid, polymerase, and envelope proteins, RT, reverse transcriptase.

Furthermore, HIV-1 can activate ERVs. Noticeably, striking similarities exist between ERV-K RT and that of HIV-1, as determined by studying the activities and crystal structure of RT enzymes with double-stranded DNA substrate ^60^. Unlike classic nonnucleoside RT inhibitors (NNRTIs), antiretroviral nucleotide analogs (NtRTIs) and phosphonoformic acid (forscarnet) inhibit ERV-K and HIV-1 RTs. 3TC-triphosphate (lamivudine-TP) exhibits ERV-K RT 40-fold higher than HIV-1 RT in the RNA-protein in-vitro assays ^60^. These findings reveal an opportunity for exploring selective inhibition of ERV-K RT enzyme in ERV RTs-positive skin cells and potentially clinical trials in psoriasis and PsD.

RNA-seq analysis on primary CD4+ T cells in culture also demonstrated that HIV-1 infection activates a set of ERV-9 LTR promoters and stimulates interferon-inducible genes with known broad antiviral activities which can suppress ERV-K, thus for example can augment ERV-directed gene expressions ^61^. Of 91 ERV-K proviruses, HIV-1 Tat protein significantly activates expressions of 26 and silences 12 variants in PBMCs from healthy donors ^62^. Another regulatory protein of HIV-1 (Rev) interacts in Trans with many ERV-K (RcRE) regulatory elements and induces nuclear cytoplasmic export and accumulation of mRNAs from ERV-K provirus in a T-lymphocytic cell line^63^.

Conceivably, HIV-1 regulatory proteins can modulate transcriptional regulatory elements or proteins of ERVs in psoriasis and psoriatic CVD. Nonetheless, different subtypes of HIV-1 cause different magnitudes of abnormal activations of ERVs. For instance, *gag-pol* regions of ERV-K are inconsistently increased in patients infected with distinct subtypes of HIV-1 ^64^. These findings imply that the regulatory proteins and different subtypes of HIV-1 can cause discordants in activation and gene expressions of ERVs, hence variably impacting the outcomes of the immune intolerance and inflammation (see further).

### Medication-induced Psoriasis

Drug-induced PsD is an age-dependent phenomenon that mostly occurred in subjects with a family history of PsD, and in those with metabolic syndrome. In this regard, the substantial role of antihypertensive drugs in PsD progression is concisely discussed below:

### Angiotensin-converting enzyme (ACE) Inhibitors

Among antihypertensive drugs, ACE inhibitors have been well-studied in the case of PsD progression, and are defined as extensively used medications for the treatment of hypertension (HTN), congestive HF, diabetes, and even the prevention of kidney diseases ^65^. According to the data analysis using the French Pharmacovigilance Database (FPVD), though psoriasis is considered one of the serious adverse events in patients receiving ACE inhibitors ^66^, the mechanism of ACE-inhibitors induced/aggravated PsD is not fully deciphered. However, it has been thought that ACE inhibitors beyond the modulation of angiotensin II (Ag II) production can also regulate the catabolism of kinins ^67^. Consequently, this class of drug augments the activity of kinins-bradykinin system, which subsequently results in the exacerbation of kinin-associated skin conditions, e.g., urticaria, angioedema, and dermatosis ^68^, following the increased levels of cutaneous inflammatory responses to allergens, and may promote the keratinocytes hyperproliferation in some skin conditions like PsD ^69^. In better words, ACE inhibitors can exert an indirect proinflammatory effect, promoting to exacerbate preexisting PsD. Two mechanisms in terms of PsD induced by ACE inhibitors are proposed: 1) an immune-dependent hypersensitivity reaction, i.e., psoriasiform eruption, and 2) a pharmacologic dose-dependent response, for example receiving ACE inhibitors in patients who are on beta-blockers (BBs) ^70^.

Previously, the implication of ACE gene polymorphism, consisting of ACE insertion/deletion (I/D) genotypes was highlighted in psoriasis vulgaris, which can contribute to both phenotypic expression and the early appearance of the disease, in part through modulation of ACE activity in degradation of bradykinin and substance P ^71^. Besides, ACE activity potentially predicts the severity of PsD, independent of ACE I/D polymorphism genotype. In comparison with control group, the psoriatic patients show higher serum levels of ACE that associates with lipid profile modulation, reflecting an increased stress oxidative status as an underlying risk factor in cardiovascular disorders ^72^. Despite disease severity and family history, the results of a meta-analysis also revealed that ID and DD genotypes may serve a protective role against psoriatic arthritis, particularly in Caucasian patients ^73^.

### Angiotensin Receptor Blockers (ARBs)

Albeit, the PsD induction/exacerbation is well-established following ARBs exposure; even so, as an adverse drug reaction (ADR) it had been underestimated and simultaneously excluded from the Summary of Product Characteristics (SmPC) of ARBs ^74^. A flared psoriatic plaque manifestation, as a cutaneous ADR which is more commonly observed by ACE inhibitors, was also detected in a female patient receiving Telmisartan ^75^, which in contrast can also significantly decrease the pro-inflammatory cytokines IL-6 and TNF-α levels ^76^. In addition, a presumable association between some rare forms of PsD such as erythema nodosum migrans (ENM) and losartan administration was observed in another patient ^77^. In contrast, losartan, exhibits a beneficial effect against imiquimod-induced psoriasis-like inflammatory response *in vivo* following topical administration (Losartan 1% ointment), more likely through the inhibition of IL17a-related inflammation mediated by suppression of the Ang II and Ang II receptor type 1 (AT1R) expression ^78^. It can be inferred that ARBs have the potential to exert dual effects in the case of PsD induction or exacerbation through different routes of administration.

### Beta Blockers (BBs)

Rising trend of BBs prescription around the world results in the increasing related side effects, particularly, dermatological reactions (such as erythrodermic psoriasis) ^79^. In this respect, the BBs can flare the preexisting PsD in one hand, and induce the *de novo* PsD in naïve individuals on the other hand. For instance, metoprolol, as a β1 selective blocker—which is one of the widely prescribed BBs in the U.S.—there are 97 reports highlighting that Ps is being the most cutaneous adverse events following metoprolol administration based on US Food and Drug Administration (FDA)’s adverse event reporting system ^80^. Beyond the exuberant pre-inflammatory responses, the modification of intracellular calcium (Ca^2+^) levels, keratinocyte proliferation, and granulocyte function by targeting cyclic adenosine monophosphate (cAMP) pathways are indicated as the underlying mechanisms involved in the BBs-induced psoriasis at the cellular level ^81^.

Of note, a recent systematic review indicated that BBs, especially nonselective drugs such as carvedilol and propranolol, may alleviate rosacea-associated facial erythema and flushing via vasoconstriction of cutaneous vessels, in part, in those with a resistant form of PsD ^82^. The latency period between drug exposure and onset of psoriatic eruption ranges from intermediate latency periods (4-12 w) for ACE inhibitors to up to one year for BBs ^83^, and six weeks to nine months for ARBs ^74,83^. Taken together, the available evidence indicates that BBs by targeting either beta 1 or beta 2 adrenergic receptors can present conflict effects in terms of PsD management.

### Calcium Channel Blockers (CCBs)

CCBs consist of dihydropyridine and non-dihydropyridine agents. Despite, the intracellular Ca^2+^ plays a key role in the keratinocyte differentiation, some studies found a significant association between PsD provocation/ exacerbation (psoriasiform eruptions) and CCBs exposure ^84^. Notably, amlodipine and atenolol for instance are also known immunomodulators to increase expression of IL-6 in experimental animals ^85^.

### Other Antihypertensive Drugs

Digoxin, a natural steroid toxin, is known as a challenging drug with a narrow therapeutic window and is mainly prescribed for congestive HF and atrial fibrillation (AF). The digoxin therapeutic effect is mediated by Na^+^/K^+^ ATPase inhibition, intracellular Ca2+ levels elevation, and subsequently leads to elevated cardiomyocytes contractility. Nevertheless, digoxin may also contribute to PsD development, ^86^. In the experimental model of psoriasis-like lesions induced by imiquimod, intraperitoneal injection of digoxin recovered the epidermal hyperplasia in the ear skin, but not the back of mice by targeting retinoid acid-related orphan receptor γt ^87^. Some case reports also represented the potential of diuretics, including furosemide, and thiazides in Psoriasis-type reactions ^88^. Noteworthy, chlorothiazide/hydrochlorothiazide exhibited a phototoxic reaction following the narrow-band ultraviolet B treatment in the psoriatic patients ^89,90^.

In **Table 3**, the details of the strengths of associations between antihypertensive drugs use and incidents of cutaneous/non-cutaneous autoimmune diseases, based on systematic reviews and meta-analyses from distinct data sources, are summarized. Strong correlations of betablockers/BB and angiotensin receptor blockers/ARBs are noticeable, while angiotensin-converting enzyme inhibitors/ACEi and calcium channel blockers/CCB exhibit low-to-moderate risks signals. These data have important implications to pharmacovigilance research, however, the causal links and the relative risk/RR ratios or absolute risk factors for these medications remained unclear. The concerns about the relevancy of the reversal of latency or activation of the ERVs in psoriatic CVD/MI manifestations were also excluded in these estimates.

**Table 3.** Antihypertensive drugs use and incidents of cutaneous autoimmune disease: strengths of associations.

| No | Study | Study Design | Sample size, Patients (n) | Condition | Drug Class | Findings | Outcome |
| --- | --- | --- | --- | --- | --- | --- | --- |
| 1 | Song et al 2021a <sup>124</sup> | Systematic review and meta-analysis | 54,509 patients | Cutaneous vulgaris | ACEi | **OR [95% CI]**<br>2.37 [1.28–4.37]<br>in Asians<br>1.52 [1.16–2.00]<br>in Europeans | A significant association between ACE inhibitors use and psoriasis incidence was found. The rate of incidents was higher among the Asian population and patients living in dry climates. |
| 2 | Song et al 2022b <sup>125</sup> | Systematic review and meta-analysis | 6, 378, 116 individuals | Cutaneous vulgaris | ACEi | 1.67 [1.31–2.13] | The associations between antihypertensive drugs and psoriasis was reported; ACE inhibitors, BBs, CCBs, and thiazide diuretics increased the risk of psoriasis. |
|  |  |  |  |  | BBs | 1.40 [1.20–1.63] |  |
|  |  |  |  |  | CCBs | 1.53 [1.23–1.89] |  |
|  |  |  |  |  | Thiazide Diuretics | 1.70 [1.40–2.06] |  |
| 3 | Song et al 2022b <sup>125</sup> | Bayesian Network meta-analysis | 5, 615, 918 individuals | Cutaneous vulgaris | ACEi | 2.09 [1.39–3.18] |  |
|  |  |  |  |  | BBs | 1.35 [0.99–1.91] |  |
|  |  |  |  |  | CCBs | 1.53 [1.07–2.24] |  |
|  |  |  |  |  | Thiazide Diuretics | 1.80 [1.23–2.66] |  |
| 4 | Ohyama et al 2020 <sup>69</sup> | FAERS database | 280,111 participants | Not mentioned | ACEi (14 types) | **ROR [95% CI]**<br>1.25 [1.14–1.37]<br>**IC [95% CI]**<br>0.31 [0.17–0.44] | Using the FAERS database, a modest association between ACEi and psoriasis was observed, suggesting that ACEi should be considered in patients who have developed psoriasis. |
| 5 | Azzouz et al 2019 <sup>66</sup> | FPVD, using the case/non-case method, between 1985 and 2018 years. | 100 reports of 770,951 ADR registered in FPVD. | Psoriasiform dermatitis, Pustular psoriasis, Erythrodermic psoriasis | ACEi (12 types) | **ROR [95% CI]**<br>2.40 [1.96–2.95] | A statistically remarkable correlation between ACEi and psoriasis indicates a potential safety signal. The outcome is acceptable following ACEi discontinuation. Therefore, Psoriasis should be mentioned in the SmPCs of all ACEi medications. This is a class-effect risk, with a time to onset < one year. |
|  |  |  |  |  | Trandolapril | 5.55 [2.72–11.33] |  |
|  |  |  |  |  | BBs | **OR [95% CI]**<br>0.94* [0.61–1.45] |  |
| 6 | Azzouz et al 2018 <sup>74</sup> | FPVD, using the case/non-case method, between 1985 and 2016 | 89 reports of 599,897 ADR registered in the FPVD | Psoriasiform dermatitis, pustular psoriasis, erythrodermic psoriasis | ARBs (7 types) | 4.86 [3.92–6.03] | Outcome was favorable (in 67% of reports) regarding psoriasis improving upon the ARB discontinuation. The case/non-case method highlighted a potential safety signal. The ADR could be a class effect, with time to onset (latency) most often < 1 year. |

Table 3 Contd.
|  |  |  |  |  |  |  |  |
| --- | --- | --- | --- | --- | --- | --- | --- |
| 7 | Logger et al 2020 <sup>82</sup> | Systematic review | 9 eligible studies | ETR, transient/persistent erythema | At least three types of BBs (Carvedilol, Nadolol, Propranolol) | ND | BBs could be an effective therapeutic option for patients with rosacea-associated facial erythema and flushing, which is resistant to conventional therapy. |
| 8 | Azzouz et al 2022 <sup>126</sup> | FPVD, using case/ non-case method), between 1985 and 2019 | 225 reports | Psoriatic lesions, psoriatic condition, psoriatic arthropathy | BBs (18 types) | **OR [95% CI]**<br>8.95 [7.75–10.33]<br>Therefore, a pharmacovigilance signal was confirmed. | The psoriasis risk with BBs is a class effect. The mean time to onset of psoriasis was 5 months and the outcome was favorable in 68% after drug discontinuation. |
| 9 | Azzouz et al 2021 <sup>127</sup> | FPVD, using the case/ non-case method, between 1985 and 2019 | 94 reports | Psoriasiform dermatitis, erythrodermic psoriasis | CCBs (11 types) | 2.45 [1.99–3.02] | There is a safety signal between CCB exposure and psoriasis risk, which is a class effect, and should be mentioned in SmPC. Latency time was < 2 years in 64% of reports. The outcome was estimated favorable in 71% of reports following CCB discontinuation. |
| 10 | Li et al 2020 <sup>128</sup> | Cohort study, population-based, TNIRD | 15, 545 patients | Not mentioned | Digoxin | **HR [95% CI]**<br>1.34* [1.04–1.72]<br>after five years of follow-up | Psoriasis risk was significantly higher among digoxin users vs. non-users after adjusting for confounders. |

**Abbreviations:** ACEi, angiotensin-converting enzyme inhibitors; ADR, adverse drug reaction; ARBs, angiotensin receptor blockers; BB, $\beta$ -blockers; CCBs, calcium-channel blockers; CI, confidence interval; ETR, erythematotelangiectatic rosacea; FPVD, French National Pharmacovigilance Database; HR, hazard ratio; IC, information component; OR, odds ratio; PPR, papulopustular rosacea; ROR, reporting odds ratio; ND, not determined; SmPC, summary of product characteristics; TNIRD, Taiwan National Health Insurance Research Database.
\* The stratified RORs/HR were significantly different ( $p < 0.05$ ).

### Meta-analysis on Antihypertensive Drugs-induced Risk of PsD

Independent risk factors may represent distinct covariates with a range of relative risk/RR ratios (low risk, intermediate risk, high risk) for the antihypertensive drugs. Our analysis of the previous studies, in accordance with class of drugs, revealed distinct characteristics, which would be grouped into the following types of inferences.

**A.** ACE inhibitors pose a higher association with psoriasis incidence, in which, in four studies conducted to assess ACE inhibitors the heterogeneity was significant (Q-value = 32.80, df= 3, I^2^ = 90.85). Moreover, the risk of PsD incident in the intervention group is 57% higher than the control group [pooled RR =1.57, 95% CI (1.89-1.95), z-value = 8.07, *p*-value < 0.001]. The forest plot represents that in five studies with the drug class other than ACE inhibitors (i.e., BBs, CBBs, and ARBs), a significant heterogeneity among studies is visible (Q-value = 375.59, df = 5, I^2^ = 98.67). In this sense, the risk of PsD in the intervention groups is 149% (more than twofold) [pooled RR =2.49, 95% CI (1.53-3.45), z-value =5.09, *p*-value < 0.001] (**Figure 3a**).
**B.** To ascertain risk stratification, the studies were divided into three categories, consisting of low, moderate, and high risk, based on PsD incidence. The risk in two studies was low, in four studies was moderate, and in five studies was high. The results revealed that the risk of psoriasis in the intervention group is 26% (in the low-risk group), 51% (in the moderate-risk group), and 197% (approximately threefold in the high-risk group) respectively, which are statistically significant (*p* <0.05; **Figure 3b**).
**C.** Among the studies included in the meta-analysis, there were five case-control (CC) studies in which the heterogeneity between studies was not significant (Q-value = 459.48, df= 4, I^2^ = 99.13). In CC studies, the risk of PsD in the intervention group was 178% (approximately threefold) [pooled RR =2.78, 95% CI (1.46-4.10), z-value =4.13, *p*-value <0.001]. Moreover, six studies were systematic reviews, in which the reported heterogeneity was not significant (Q-value = 5.59, df = 5, I^2^ = 10.62). The risk of PsD occurrence in this type of studies was 47%, which was statistically significant [pooled RR =1.47, 95% CI (1.36-1.58), z-value =26.00, *p*-value <0.001] (**Figure 3c**).
**D.** Regarding psoriasis types, in five studies cutaneous/vulgaris type with non-significant heterogeneity was reported (Q-value = 359.48, df = 4, I^2^ = 0.99.13). However, the forest plot drawing depicts that the risk of cutaneous/vulgaris PsD in the intervention group is 178% (more than threefold) [pooled RR=2.78, 95% CI (1.46-4.10), z-value = 4.13, *p*-value <0.001] (**Figure 2d**).
**E.** Furthermore, we performed a subgroup analysis to realize the risk of each class of drugs in PsD development. Considering a significant heterogeneity (*p*-value <0.05), the risk of disease in the ACEi users was 62% higher than that of individuals who did not use it. This analysis shows that the relative risks of PsD incident in the users of BBs, CCBs, ARBs, and thiazides was 3.16, 1.78, 3.50, and 1.58 times higher, respectively, than those who did not use these drugs (**Figure 3e**).
**F.** These findings expand previous estimates and show that significant risk differences exist between ACEi and other antihypertensive classes, in which the magnitude of risk for the analyzed agents is in order of ARBs > BBs > CCBs > ACEi > Thiazides (**Figure 3f**). Whether the underlying cellular/molecular mechanisms for potentials of the high-risk ERV mutants and medications that plausibly impair target cells may be reciprocally exclusive, and together might trigger immune intolerance and inflammatory response (**Figure 4**).

**Figure 3.**
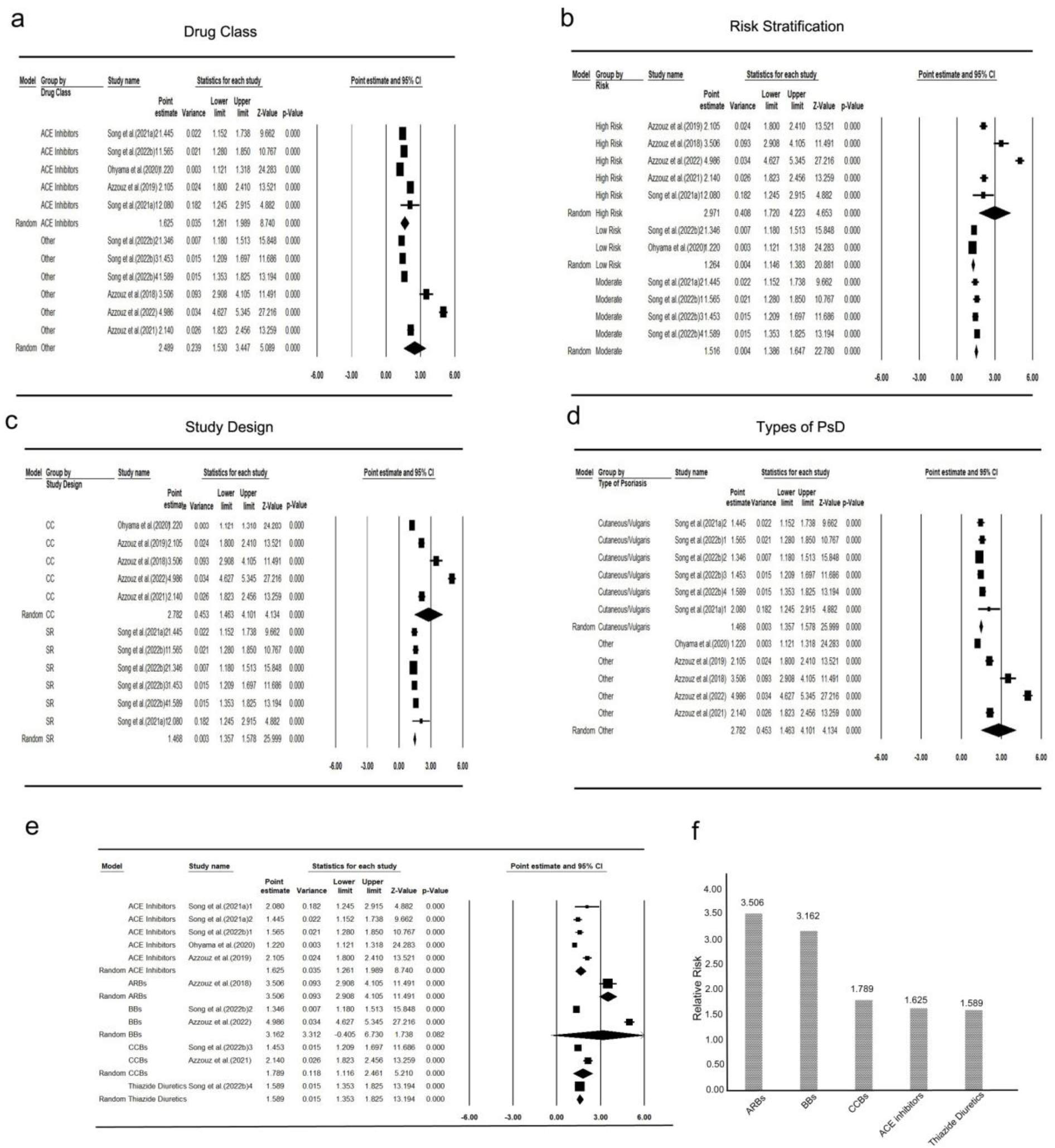
The Comprehensive meta-analysis of studies assessed antihypertensive drugs-induced psoriasis. **a,** the forest plots of pooled RR (CI 95%) estimations for distinct classes of antihypertensive drugs, **b,** the risk stratification includes low, moderate, and high-risk rates. According to the risk of PsD incidence reported in various studies, we defined low risk as RR < 1.35, for moderate risk = 1.4 to 2.0, and high risk: 2 > RR. **c,** pooled RR measured in accordance with study design (CC and SR), **d,** types of PsD (cutaneous vulgaris, other types), **e,** the subgroup analysis to determine the risk of each drug class in PsD development, **f,** the probable risk of psoriasis induction derived from antihypertensive drugs. ***Abbreviations*:** CC, case-control; RR, relative risk; SR, systematic review

**Figure 4.**
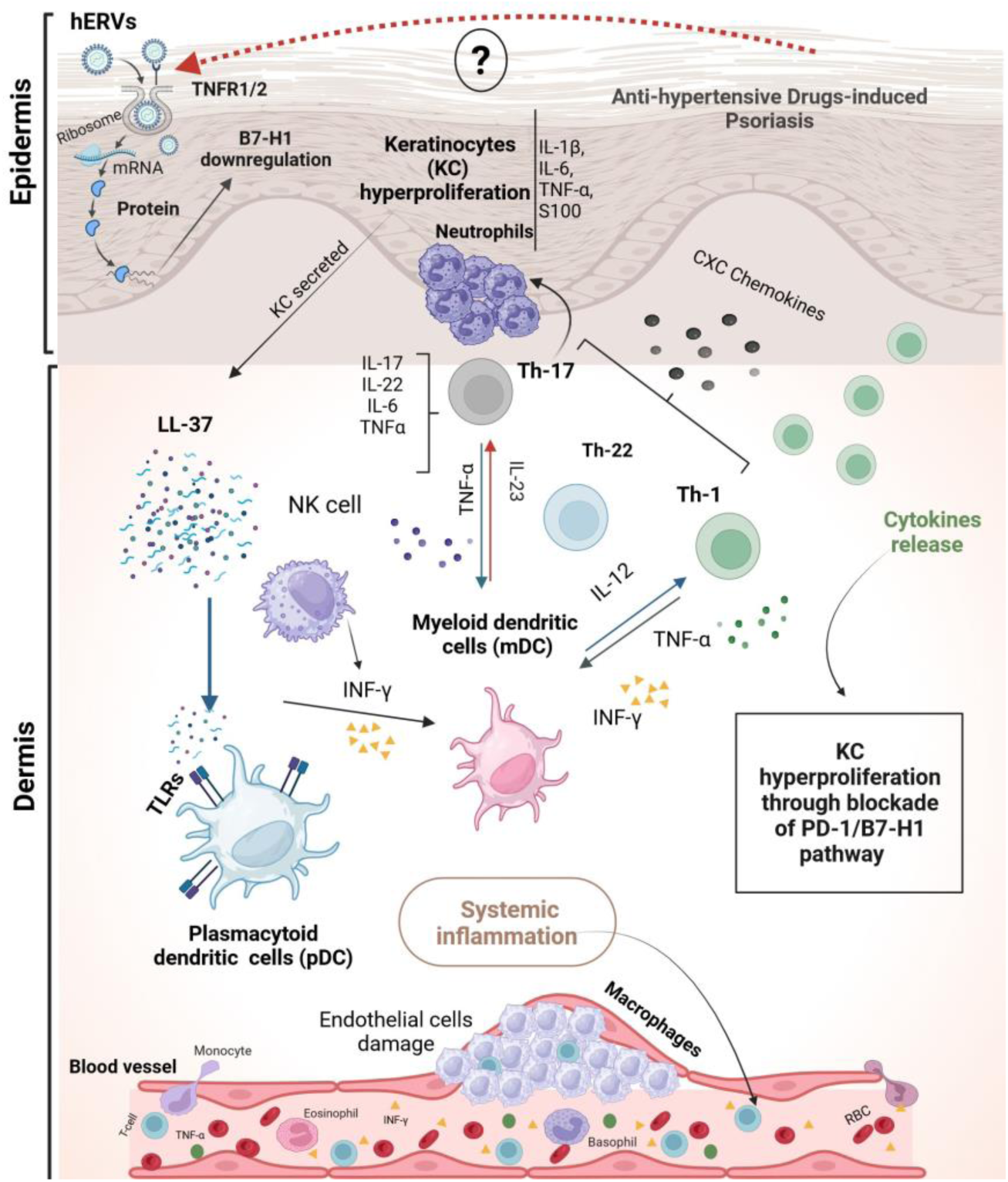
The schematic illustration of drug-induced psoriasis pathogenesis, and possible hERVs role in psoriatic CVD. The mechanism of psoriasis induction/exacerbation initiates through KC hyperproliferation following KC exposure to a variety of stimuli (e.g., genetics, infections, drugs, environment factors, UV irritation), resulting in neutrophil degranulation (attracted by CXC chemokines), and LL-37 secretion, followed by the cytokines overexpression, e.g., IL-1β, IL-6, TNF-α, and S100 (as a cytosolic Ca^2+^-binding protein involved in the immune response and inflammatory events) from respective cells and activation of the inflammatory responses. In the next step, KC-secreted LL-37s bind to the TLRs to activate both dermal DCs (pDC, mDC). The activation of Th1 and Th17 is also mediated by IL-23, and then IL-17 and IL-22 are released from Th17, while some important cytokines such as TNF-α and INF-γ released from Th1 to perpetuate the KCs proliferation. Upon the psoriasis plaque build-up in the epidermis/dermis (skin lesion) and subsequent ECs dysfunction, the levels of circulating proinflammatory cytokines also increase and, in turn, can infiltrate to the blood flow to cause systemic inflammation. ***Abbreviations***: CVD, cardiovascular disease; CXC chemokines, C-X-C motif chemokines; EC, endothelial cell; hERVs, human endogenous retroviruses; INF-γ; interferon gamma; KC, keratinocyte; LL-37, cathelicidin, 37-residue peptide with a pair of leucines (LL) at the N-terminus; mDC, myeloid dendritic cell; pDC, plasmacytoid dendritic cell; Th, T-cell helper; TLR, toll-like receptor; TNF-α, tumor necrosis factor-alpha; TNFR1/2, TNF-receptor 1/2

## Discussion

Overwhelming evidences support the notion that ERV-derived elements (-E, -K, -W) are involved in the pathophysiology of multiple autoimmune diseases, through instigation of human innate immunity, and the latter mechanism is referred to the activation of specific signaling mechanisms, i.e., TLR-dependent pathways, through ERV-related RNAs and/or proteins ^91^. Of these, aberrant expression profiles of the ERV-K (HML-2), considering polymorphic ERV loci (such as ERV-K dUTPase for psoriasis susceptibility 1 locus) ^92^ appear to be of a promising diagnostic and prognostic value in the pathogenicity of various diseases ^93^.

However, conflicting datasets are reported in regard to expressions of different ERVs (E, K, W, 09) in psoriasis (**Table 1**, **2**). These variants are clearly distinct in terms of the types of mutations, transcriptional and gene expression activities. This is particularly evident in relations to differences in stable viral transcripts/mRNA levels and structures in different autoimmune diseases ^3,94^, compared to psoriasis (**Table 2**). Notably, the expression of ERV-E envelope glycoprotein is active in lesional cells, but is suppressed in PBMCs presumably as a result of an antiviral defense mechanism ^13^, or its unessential role in normal functions of immune cells in circulation. Although anti-ERV/RT activities are compromised in epidermal skin cells in psoriasis, both Env and RT proteins might serve as autoantigens in psoriasis.

ERV-E antigens seem relevant as their elevated levels positively correlate with psoriatic pathology, and might be clinically useful as surrogate diagnostic antigens. Whether ERV-E Env and retroviral RT proteins (in the absence of viral particles) in lesional cells are engaged with other viral retroelements or transcription and aberrant differentiation of keratinocytes, or simply engaged as autoantigens in triggering inflammation in psoriasis, are unclear. In addition, expressions of ERV-K and ERV-9 genes are lower in lesional psoriasis ^20^, conflicting earlier reports that ERV-09 activity may be relevant to the disease pathology (**Table 1**). Further work is needed to determine consistency (interassay, intraassay) and discrepancies, including utility of ERV genes as diagnostic and prognostic markers, with emphases on reliability, sensitivity and specificity of these biomarkers in relations to different ERVs strains and cohorts.

Plausible breakdown of cell-cycle checkpoint in keratinocytes by ERVs are also unexplored in psoriasis. Multiple mutants of ERV-K trigger Th1/Th17 response among T lymphocytes that result in increased release of cytokines (TGF-α, NF-κB, IFN-γ, and TNF-α) as a consequence of triggering Th1/T17 response among T lymphocytes in psoriasis (**Table 2**). These cytokines appear to encounter uncontrolled proliferation and differentiation of keratinocytes by unleashing danger signals that in turn can fuel ERVs from within genome structures in dermal CD3+/CD4+ T-cells infiltrates.

According to the recent immunosenescence paradigm, a negative lipid body (LB^-^) of proinflammatory foamy macrophage (CD14+CD16+) and trained inner immunity, deriving atherosclerosis-cardiovascular disease, are more likely involved in ERV-K induction ^95^. These particles are engaged as the protective foamy virus, promoting autoimmunity, particularly in facing with intracellular pathogens, toxins, and/or various tumors ^95^. In the clinical setting, pulmonary arterial hypertension (PAH) is defined an idiopathic vascular dysfunction, resulting in heart failure and death. Recently, it has been well-documented that inflammatory response and autoimmunity are jointly implicated in the PAH development, in which the ERV-K emerges a crucial role ^96^. In this regard, the lung tissue of the patients with PAH exhibits a higher load of ERV-K envelopes, as well as dUTPase mRNA mainly expressed in lung CD68+ macrophages, compared with healthy counterpart ^96,97^. Further research is immensely required to shed a new light on presumable link between ERV-K and psoriasis-induced inflammatory heart diseases.

Methylation of ERV mRNAs is another layer of transcriptional regulation at works that might be dysregulated in keratinocytes. In support of this plausibility, RNA methylation reportedly maintains cellular integrity by clearing reactive ERV-derived RNA species ^98^ in addition to the transcriptional silencing of the LTRs of certain ERVs by DNA methylation ^4^. This would be particularly relevant when transcriptional silencing of the LTR-like promoter becomes less stringent or compromised by either genomic mutations or exogenous risk cofactors like the HIV-1 trans-complementation of ERVs. Mutated ERVs that are differentially expressed may be more susceptible to activations by exogenous retroviruses than other autoimmune stimuli. Our assessments support research considerations to determine whether HIV-1 subtypes alter expressions of ERV-K/-9 directed stable mRNAs to the extent that variably exacerbates the disease outcomes.

As for the discrepancies in incidences of psoriasis and PsD, apart from confounding variables in different demographic/ethnicity measures, covariates (e.g., in RNA-seq and epitope mapping assays) can compromise the accuracies/reliabilities of the outcome measures. Importantly, as the ERV RTs expressions can be verified by laboratory tests, it would be interesting to determine whether commercially available NtRTIs ameliorate symptoms of psoriasis and PsD in persons living with or without HIV-1.

A recent Mendelian randomization analysis enriched for non-MHC markers independent of the other risk factors indicates that the chronic systemic inflammation in the coronary artery disease/CAD causes psoriasis (OR=1.11, *p*=3×10^-6^) ^10^. This is a weak association, considering the larger sample sizes, and excluded the MHC signals and tissue-driven inflammation. Others assert that patients with psoriasis are at a higher risk of CVD ^99^ and acute myocardial infarction/MI (HR=2.1, 1.27-3.43, *p*=0.004) ^100^. The etiology of higher risk CVD/MI events in psoriatic patients is controversial, in part because most such analyses are based on insufficient risk assessments or unadjusted confounding variables.

While response rates for biologics treatment such as anti-IL-17A monoclonal antibody targeting proinflammatory cytokine is reportedly near 68% at 5 years for psoriasis and psoriatic arthritis ^101^, severity and paucity of remission with or without cardiovascular involvements and other disease burdens such as infections with HIV-1 retroviruses remain a real concern. For instance, the use of systemic immunosuppressive drugs is contraindicated while administering biologics drugs long-term is often undesirable primarily because of suboptimal efficacy and high adverse events.

Furthermore, treatment modifications, such as dose reduction, dose escalation cessation and restart of treatment, concurrently or in conjunction with systemic treatment options, such as methotrexate are recommended and often necessary for a balanced evaluations of the clinical outcomes and response rates ^102^. In contrast to Steven Johnson Syndrome/SJS or other drug eruptions, which are largely reversible or mitigated upon cessation or modification of the causative drugs ^103,104^, to gain a complete remission in psoriasis and reducing the disease burden in psoriatic CVD remains challenging.

Notably, inhibition of ERVs expression by antiretroviral drugs exhibit a dual beneficial and negative effects in a microbiota-associated psoriasis model in mice, topically challenged with bacteria. A combination of nucleotide and nucleoside RT inhibitors, tenofovir and emtricitabine, impairs ERVs-induced T-cell responses, but it also weakens the dermal tissue repair functions ^105^. These findings are modestly comparable to the highly prevalent human psoriasis, psoriasis vulgaris or plaque psoriasis, but are relevant to bacterial infections-associated psoriasis. Of interest, the authors’ findings show that skin inflammation is associated with increased expression of ERVs, which can be suppressed by antiretroviral drugs. Apart from considerable differences in murine and human ERVs, including ERV genes sequences and locations, distinct immunological responses have been observed. Unlike in rodents, endogenous viral particles are undetected in lesional and nonlesional tissues in human psoriasis. In murine model, ERVs in association with skin microbiota promote type I interferon (IFN-I) similar to intestinal microbiota ^105^, whereas human mutated ERVs increase interferon gamma/IFN-γ ^59^ (**Table 2**).

To the best of our knowledge, the causes of inflammation and increased susceptibility to CVD and risk of MI in relations to early and recent antiviral regimens ^106,107^, apart from HIV-1 infection per se ^108,109^—independent of dyslipidemia and lifestyle risk factors—and ERVs gene expressions signatures are previously unaccounted for, even though most current therapies are less toxic and more efficacious than earlier antiviral agents ^110^. In a recent commentary in New England Journal of Medicine, the author is concerned about insufficiency of the evidence for mitigating CVD risk in persons living with HIV-1 ^109^—despite considering atherosclerotic risk factors and even effective antiretroviral therapy—which is relegated to because of an underestimation of the cardiovascular risk assessment using a traditional disease diagnostic algorithm ^111,112^.

Although a distinct guideline is lacking to utilize in these conditions, the likelihood of h-ERV dysregulation induced by HIV-1 might also be deriving the increased risk of CVD with or without psoriatic manifestations (**Figures 4**, **5**).

**Figure 5.**
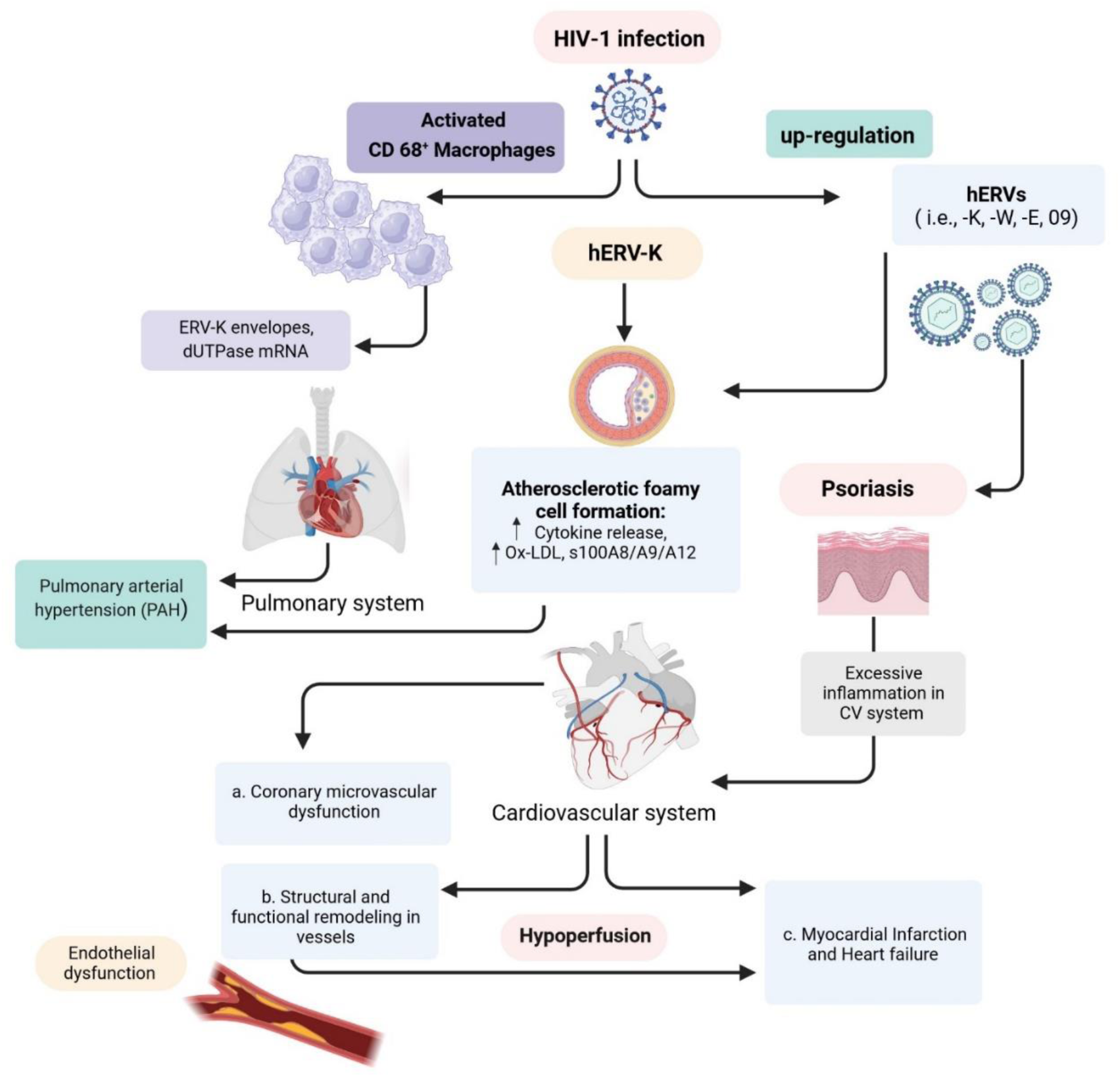
HIV-1-induced hERV up-regulation and possible cardiovascular effects. Upon cell entry and expression, human ERVs may modulate TNFR1/2 (AdR), to downregulate the B7-H1 expression, which is expressed in normal KC. Of hERVs subtypes, the higher load of hERV-K envelops is presumably involved in some CVDs pathogenesis, such as PAH, through promoting the atherogenesis process. Upregulation of hERVs expression can also occur following exogenous HIV-1 infection. The CD68+ macrophages are mainly participated in this process, where the dUTPase mRNA is highly expressed. However, the role of hERVs in hypertensive drugs-induced psoriasis remains to be determined. ***Abbreviations*:** CV, cardiovascular; hERVs, human endogenous retroviruses; HIV-1, human immunodeficiency virus-1; Ox-LDL, oxidized low-density lipoprotein

Among drugs induced PsD, ACE inhibitors can exert an indirect proinflammatory effect, promoting PsD exacerbation. In this line, two mechanisms were proposed: 1) an immune-dependent hypersensitivity reaction, i.e., psoriasiform eruption, and 2) a pharmacologic dose-dependent response, for example receiving ACE inhibitors in patients who are on beta-blockers (BBs) ^70^. Previously, the implication of ACE gene polymorphism, consisting of ACE insertion/deletion (I/D) genotypes was highlighted in psoriasis vulgaris, which can contribute to both phenotypic expression and the early appearance of the disease, in part through modulation of ACE activity in degradation of bradykinin and substance P ^71^. Besides, ACE activity have potential to predict the PsD severity, independent of ACE I/D polymorphism genotype. In the clinical settings, the psoriatic patients showed higher serum levels of ACE that had association with lipid profile modulation, reflecting an increased stress oxidative status, as an underlying risk factor in CVD ^72^.

Of note, the latency period between drug exposure and psoriatic eruption varies among different drugs. Intermediate latency periods (4-12 w) were observed for ACE inhibitors, while longer latency periods (up to one year) for BBs ^83^, and ARBs between 6 weeks and 9 months ^74,83^. Taken together, the evidence suggests that BBs—by targeting either beta 1 or beta 2 adrenergic receptors—can present conflict effects in terms of PsD management. ERVs presumably trigger autoimmune psoriasis and are inducible by HIV-1. It is tempting to speculate that some medications on the other hand simultaneously could promote activations of the endogenous and nonendogenous retroviral gene expressions in keratinocytes and endothelial cells, in part through induction of proinflammatory molecules in psoriatic CVD (**Figure 4**, **5**).

### Research Limitations

Previous studies on mutated ERV expressions suffer from several drawbacks. Inferences were made from the analyses of the cDNAs prepared from total RNAs extracted from whole tissue, liquid/solid biopsies for differential display and mapping studies. Yet, precise quantifications of mRNAs are challenging, for example because of variabilities in transcription and degradation rates, and partial expressions. These observations also lack visualization of cell-to-cell variability and none of the previous methods provide subcellular resolution.

Others utilized conventional in-situ hybridization or immune staining methods that exclude diffraction limited fluorescent proteins. Such studies are based on limited thin-cell layer tissue sections with a low multiplexing capacity and exclude poly-A truncated, untranslated and noncoding RNAs. These methods would restrict capturing multiple transcriptional signals related to ERVs expressions and the intracellular communications such as enhancer-promoter interactions, as well as post-transcriptional processes, proteins syntheses and trafficking, and cell-to-cell interaction signals in the lesional compared to healthy tissues.

An abundance of mutations in dUTPase ^8,92^ may also hinder the enzyme activity and overload the DNA mismatch repair system. The genetic heterogeneity—mutational numbers and burden in the lesions—are challenging to stratify. Although it appears useful, the results are insufficiently addressed in various types of psoriasis and PsD. For an accurate/unbiased representation and interpretation of the transcriptional landscape of the candidate mutants, for instance ERV-K ORFs ^8^ and noncoding LTR-like repetitive sequences or duplicate genes ^7^, the panel of mutated ERVs should be aligned with the ‘pangenome’ database nucleic acid sequences, currently includes genome sequences of 47 people ^113^ for an integrated analysis of the biomarkers.

Whether the aberrant expressions of ERVs, chemokines release and loss-of-functions of the mutant proteins impair the normal quiescence in cell cycles are independent of the telomere proteins and telomerase reverse transcriptase (TERT) activity are unaddressed. Experimental considerations are needed to validate the mutated ERVs by identifying such molecular circuit connections, apart from other covariates, for instance, inherent to RNA-seq outcome measures and assessments of the confounding variables in clinical investigations involving ERVs. It should be mentioned that a heterologous set of ERVs can also be transmitted via organ transplants or blood transfusions that may in part account for a transient activation or suppressions of hybrid ERV genes.

The biological plausibility based on the clinical and basic science principles for the mutated ERVs as the necessary causes of psoriasis and psoriatic CVD remains questionable. Odds ratios/ORs or strengths-of-associations are insufficient to support the ERVs and certain classes of the antihypertensive drugs, or cell-specific metabolic risk factors, as direct or indirect causes of psoriasis and PsD. Despite the noticeably high variations between different studies, some antihypertensive drugs indicate a considerably impressive magnitude of associations or higher ORs than others. Assuming that cutaneous inflammatory disease and psoriasis are rare adverse events associated with antihypertensive drugs, the unadjusted ORs can overestimate the actual relative risk values.

As the first prospective cohort study (1996-2008), published in the JAMA dermatology in 2014, to find a rational link between hypertension/antihypertensive drugs and PsD induction based on data gathered from the Nurses’ Health Study (NHS) of women, the authors indicated that long-term (≥ 6 years) hypertensive status and BBs regularly use, but not other hypertensive drugs, were highly associated with the PsD progression [HR=1.27, 95% confidence interval (CI), 1.03–1.57] and 1.39 (95% CI, 1.11–1.73), respectively] ^114^.

Some antihypertensive medications exhibit immunomodulatory effects that can modulate the levels of proinflammatory proteins. For instance, the beta blocker, atenolol, and CCB amlodipine can increase IL-6 levels ^85^, probably through activating the respective gene promoters ^115^. In contrast, certain ACE inhibitors for instance are known to downmodulate cytokines activity and cytokine-mediated cell growth in PBMCs ^116^, presumably independent of ACE. However, it is unlikely that ACEi and ERVs would increase the risks of PsD incidents via an additive/synergist shared proinflammatory immune response. Whether or not the ACEi drugs can influence off-target (transcriptions, epigenetics, or proteins) modifications in ERV expressions has yet to be determined.

Often, one needs to go beyond the data to attain reasonable inferences without making unwarranted assumptions. The consistency-of-associations in the ORs—backed by the empirical data shown by the quantity (percentage) values—nevertheless demonstrate the ‘high-risk’ ERV mutations plausibly in connection with RRs estimated for medications associated PsD, could be theoretically exclusive risk factors. Considering that psoriatic disease/PsD is the common denominator, covariates are highly likely between the ERVs expressions and such risk cofactors. While the latter seem supportive or contributory causes, the former may be necessary and sufficient causes of psoriasis and psoriatic cardiovascular disease. These conditions may interlink and triggered by activations of the mutated ERVs.

### Future Directions

A useful approach for exploring causality that is currently unaddressed by genome-wide association studies for numerous ERV variants ^7^ may be afforded by advanced single-cell RNA sequencing (scRNA-Seq) technology ^117^. Advance multiplex mRNA in-situ hybridization and imaging techniques can be applied to perform targeted observations to quantify accurate counts of ERV gene expressions. Complementary techniques combining hybridization-based single-cell sequencing and spatially resolved transcriptomics linked to computational tools would be better to understand cell-to-cell interactions in complex tissue biopsies ^118–120^. A promising recent approach shows prefeasibility results from a noninvasive high-resolution technique that captures RNA profiling of full-thickness lesional and nonlesional biopsies from the same psoriatic patient ^121^.

Such techniques could be applied to explore ERVs profiling simultaneously with different biomarkers in early diagnosis and treatment responses in inflammatory skin conditions in the context of whole tissue biopsies. These novel approaches would allow simultaneously quantify and map ERV variants in the two-dimensional lesioned tissues, and enable visualizing single-cell transcriptomics and their spatial cellular localization with improved sensitivity and reliability—to overcome limitations of the previous studies and inferences made for interactions of the diseased keratinocytes or endothelial cells with the status of immune cells in inflammation.

A useful approach for exploring causality that is currently unaddressed by genome-wide association studies for numerous ERV variants ^7^ may be afforded by advanced single-cell RNA sequencing (scRNA-Seq) technology ^117^. Advance multiplex mRNA in-situ hybridization and imaging techniques can be applied to perform targeted observations to quantify accurate counts of ERV gene expressions. Complementary techniques combining hybridization-based single-cell sequencing and spatially resolved transcriptomics linked to computational tools would be better to understand cell-to-cell interactions in complex tissue biopsies ^118–120^. A promising recent approach shows prefeasibility results from a noninvasive high-resolution technique that captures RNA profiling of full-thickness lesional and nonlesional biopsies from the same psoriatic patient ^121^.

Such techniques could be applied to explore ERVs profiling simultaneously with different biomarkers in early diagnosis and treatment responses in inflammatory skin conditions in the context of whole-tissue biopsies. These novel approaches would allow simultaneous quantification and map ERV variants in the two-dimensional lesioned tissues, and enable visualization of single-cell transcriptomics and their spatial cellular localization with improved sensitivity and reliability—to overcome limitations of the previous studies and inferences made for interactions of the diseased keratinocytes or endothelial cells with the status of immune cells in inflammation.

In addition to targeting mutated ERVs in keratinocytes’ progenitor cells, and perhaps even in endothelial cells, possibly using gene medicines such as CRISPER-guided therapies targeting mutations ^122^, future therapies could also consider reprogramming progenitor cells by means of tissue microbiome modifications. To this end, alterations in the skin and intestinal microbiome play a critical role in psoriasis ^123^. Such interventions might restore the normal proliferation and differentiation of the keratinocytes and endothelial cells, without compromising the normal functions of the T-cells, and dendritic-like cells in recycling cellular debris.

A better understanding of the specific ‘high risk’ candidate ERVs gene variants would be afforded in relations to major histocompatibility genes/MHC clusters of genes that code proteins on keratinocyte cell surface. Results from such research can be analyzed in relation to immune-based functional studies at the RNA-protein structure-function and antigenicity studies. Further research should be optimized to differentiate shared haplotype roles in disease-specific root causes, where key immunomodulatory biomarkers/cytokines are often perceived (and clinically utilized in the diagnosis of psoriasis) as sole surrogate markers for outcome measures in clinical investigations as well as risk evaluations and mitigations strategy studies.

Assuming that the controlled study sample size is sufficient (e.g., **Table 2**, **3**), future studies should represent predefined cohorts, for instance, stratified cohorts with active disease to mild, severe psoriasis and those in remission, as well as medication-untreated special populations, such as psoriatic CVD, e.g., coronary artery disease, to dissect confounding variables. Such research can optimize signals-to-noise ratios for dependent variables and determine what elements in fact trigger and aggravate psoriasis and psoriatic CVD.

Given the dichotomous potential connections between h-ERVs activations and the antihypertensive drugs on one hand, and the epigenetics and/or genetic polymorphisms on the other hand, the need for further clinical studies is highly reasonable in PsD patients.

Covariations may exist between the mutated ERVs expressions in keratinocytes, immune and endothelial cells, and their interactions with medications-induced/aggravated psoriasis and psoriatic CVD. Noteworthy, this appears to prime a great demand to bridge the gap between medication-induced/aggravated PsD and the possible role of ERVs using cutting-edge modalities in future research.

These assessments have important clinical implications. To elucidate the roles of ERV variants in psoriasis and intertwined trans-disease complications, incidents of psoriatic CVD, and medication-induced psoriasis, future research should incorporate longitudinal samples from liquid/tissue biopsies. For instance, from vascular endothelia and cardiomyocytes, based on predefined inclusion criteria, ultimately toward developing genetic precision medicine-based diagnostic tests and medical treatments. Developing a risk stratification based on ERVs expressions and high-risk antihypertensive drugs in stratified samples may be useful for better evaluations of the responders and nonresponders to the therapies. Considering that the utilities of the inflammation-targeted medications have nearly been exhausted, the development of cell-specific diagnostics and gene medicines, as well as NtRTIs, appear sensible next steps to attain stable remissions without compromising safety.

## Conclusion

Accumulating data suggests that there is a direct line between expressions of human ERVs and immune intolerance. Evidence is scant to prove that a discrete set of disease-specific ERV mutations cause autoimmune psoriasis and psoriatic cardiovascular disease/CVD. However, unanticipated high expressions of the ERV genes—such as reverse transcriptase/RT and envelope/Env proteins and differentially expressed transcripts containing a variety of point mutations or deletions found in lesional cells—indicate that involvements of ERVs in triggering continuous proliferation and aberrant differentiation of keratinocytes are highly likely, which in turn release cellular autoantigens and ensuing inflammation. Medications-induced activations of ERVs LTR-like promoters in keratinocytes and vascular endothelial cells remain questionable. Notably, suboptimal efficacies of the available drugs for psoriasis—from steroids to biologics targeting interleukins to TNF-α inhibitors—may be because of overexpression of the mutated ERVs in keratinocytes and detrimental effects of proinflammatory polarization in affected tissues. While steroid therapy temporarily relieves inflammation, but it would also compromise the capacity of the immune cells against clearance of the autoantigenic cellular debris shedding from keratinocytes in the affected tissues. Further studies are needed to determine whether covariations in drugs-induced psoriasis are modulated by mutated ERVs or vice versa and explore novel therapies such as nucleotide RT inhibitors (NtRTIs) and advanced gene-expression-modifying drugs to disable or reverse expressions of mutated ERVs.

## Disclosure

### Author Contributions

A.S. – reviewing/writing, psoriatic cardiovascular disease, medication-induced psoriasis, table 3, figures 1, 4, 5. H.H. – statistical analysis, figure 3. M.R.S. – initiated the study, conceptualization, and preparation of the initial draft, table-of-contents, abstract, introduction, ERVs/HIV-1 review/writing, tables 1, 2, Figure 2. A.S., M.H.M., M.R.S. – discussion and editing. The authors conducted their reviews independently. Figures 4 and 5 were created with BioRender.

## Data Availability

All data produced in the present study are available upon reasonable request to the authors

